# KL-MOB: Automated Covid-19 Recognition Using a Novel Approach Based on Image Enhancement and a Modified MobileNet CNN

**DOI:** 10.1101/2021.05.13.21257164

**Authors:** Mundher Mohammed Taresh, Ningbo Zhu, Talal Ahmed Ali Ali, Mohammed Alghaili, Asaad Shakir Hameed, Modhi Lafta Mutar

## Abstract

The emergence of the novel coronavirus pneumonia (Covid-19) pandemic at the end of 2019 led to chaos worldwide. The world breathed a sigh of relief when some countries announced that they had obtained the appropriate vaccine and gradually began to distribute it. Nevertheless, the emergence of another wave of this disease has returned us to the starting point. At present, early detection of infected cases has been the paramount concern of both specialists and health researchers. This paper aims to detect infected patients through chest x-ray images. The large dataset available online for Covid-19 (COVIDx) was used in this research. The dataset consists of 2,128 x-ray images of Covid-19 cases, 8,066 normal cases, and 5,575 cases of pneumonia. A hybrid algorithm was applied to improve image quality before conducting the neural network training process. This algorithm consisted of combining two different noise reduction filters in the images, followed by a contrast enhancement algorithm. In this paper, for Covid-19 detection, a novel convolution neural network (CNN) architecture, KL-MOB (Covid-19 detection network based on MobileNet structure), was proposed. KL-MOB performance was boosted by adding the Kullback–Leibler (KL) divergence loss function at the end when trained from scratch. The Kullback–Leibler (KL) divergence loss function was adopted as content-based image retrieval and fine-grained classification to improve the quality of image representation. This paper yielded impressive results, overall benchmark accuracy, sensitivity, specificity, and precision of 98.7%, 98.32%, 98.82%, and 98.37%, respectively. The promising results in this research may enable other researchers to develop modern and innovative methods to aid specialists. The tremendous potential of the method proposed in this research can also be utilized to detect Covid-19 quickly and safely in patients throughout the world.

## INTRODUCTION

The novel coronavirus 2019 (Covid-19) is a recently recognized disease caused by severe acute respiratory syndrome coronavirus 2 (SARS-CoV-2). Being highly transmissible and life-threatening, it has rapidly turned into a global pandemic, affecting worldwide health and well-being. Tragically, no effective treatment has yet been approved for patients with Covid-19. But the patient can have a good chance of survival if they are diagnosed early enough.

As a widely available, time- and cost-effective diagnosing tool, Chest X-ray (CXR) can potentially be used for the early recognition of Covid-19. Nevertheless, Covid-19 can share similar radiographic features with other types of pneumonia, making it difficult for radiologists to distinguish manually. With such a difficulty, manual detection of Covid-19 becomes time-consuming and mistake-prone as it is left to the intuitive judgment of the radiologist. As such, it is highly recommended to adopt automated detection techniques.

With the rapid spread of Covid-19 globally, researchers have begun using state-of-the-art DL techniques for the automated recognition of Covid-19. The initial lack of Covid-19 data compelled earlier research to use pre-trained networks to build their own models (Narin et al., 2020; Ozturk et al., 2020; Apostolopoulos and Mpesiana, 2020; Civit-Masot et al., 2020; Albahli, 2020; Sethy and Behera, 2020; Apostolopoulos et al., 2020; Chowdhury et al., 2020; Punn and Agarwal, 2020; Farooq and Hafeez, 2020; Maghdid et al., 2020; Hemdan et al., 2020). Just a few months after being discovered, Covid-19 had infected millions of peoples worldwide. Consequently, a mid-range dataset of positive cases has been made available for public use, which was uploaded from https://github.com/lindawangg/COVID-Net/blob/master/docs/COVIDx.md by (Wang et al., 2020). This, in turn, has enabled further progress in developing new, accurate, in-depth models for Covid-19 recognition (Ahmed et al., 2020; Afshar et al., 2020; Ucar and Korkmaz, 2020; Luz et al., 2020; Hirano et al., 2020; Rezaul Karim et al., 2020). However, some medical imaging issues usually pose difficulties in the recognition task, reducing the performance of these models. These issues include, but are not limited to, insufficiency of training data, inter-class ambiguity, intra-class variation, and visible noise. These problems indeed necessitate a significant enhancement of the discrimination capability of the associated model.

One way around these issues is to utilize proper image-preprocessing techniques for noise elimination and contrast enhancement. A close look at the available images reveals the presence of various types of noise, such as impulsive, Poison, speckle, and Gaussian noise. See Figure 1 (the most common types of noise in X-ray images (Paul et al., 2018)). However, the most prevalent studies have been dedicated to only some of these types of noise, e.g., Gaussian and Poison. In particular, among many other techniques, histogram equalization (HE)) (Civit-Masot et al., 2020; Tartaglione et al., 2020), adaptive total variation method (Punn and Agarwal, 2020), Contrast Limited Adaptive Histogram Equalization (CLAHE) (El-bana et al., 2020; Saiz and Barandiaran, 2020; Maguolo and Nanni, 2020), white balance followed by (Siddhartha and Santra, 2020), intensity normalization followed by CLAHE (Horry et al., 2020; El Asnaoui and Chawki, 2020), histogram equalization (HE), Perona-Malik filter (PMF), unsharp masking (Rezaul Karim et al., 2020), and Gaussian filter (Jamil et al., 2020) are, as far as we are aware, the only adopted techniques in Covid-19 recognition to date. Moreover, the utilized filters can result in blurry (by Gaussian filter) or blocky (by PMF) features in the processed image. Accordingly, there is still room to incorporate more effective preprocessing techniques to further increase the system’s accuracy.

**Figure 1.**
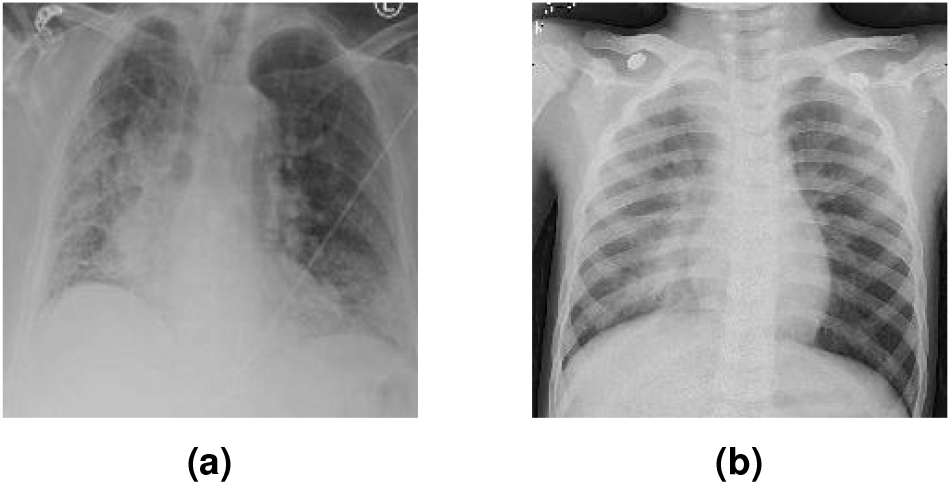
Noisy images: a) image with impulsive noise and b) image with Gaussian noise.

Motivated by the outstanding results in the previously mentioned works as well as the need for close-to-perfect recognition models, this paper integrates novel image-preprocessing enhancement with deep learning to meet the challenges arising from data deficiency and complexity. Specifically, we combine adaptive median filter (AMF) and Non-Local Means filter (NLMF) to remove the noise from the images. Indeed, many works have analyzed the performance of these two filters in X-ray imagery denoising, e.g., (Kim et al., 2020; Raj and Venkateswarlu, 2012; Rabbouch et al., 2020; Sawant et al., 1999; Mirzabagheri, 2017), demonstrating their superiority to various filters including the ones in the cited works in terms of removing impulsive, Poison, and speckle noise while preserving the useful image details. We then utilize the CLAHE approach to enhance the contrast of the denoised images. The enhanced images are finally fed into a Mobile CNN for training and validation phases. The motivation behind choosing Mobile CNN is that it not only helps to reduce overfitting but also runs faster than regular CNN with many fewer parameters (Howard et al., 2017; Yu et al., 2020). Inspired by (Alfasly et al., 2019; Alghaili et al., 2020) we adopt the KL divergence loss to measure how far we are from the optimal solution during the iterations. We evaluated the performance of the proposed framework on the COVIDx dataset in terms of a wide variety of metrics: accuracy, sensitivity, specificity, F1-score, area under the curve, and computational efficiency. Simulation results reveal that the proposed framework significantly outperforms state-of-the-art models from both quantitative and qualitative perspectives. The main contributions of this work can be summarized as follows:

- We propose an automated end-to-end deep learning framework based on MobileNet CNN with KL divergence loss function for Covid-19 recognition.
- We incorporate a novel preprocessing enhancement technique consisting of AMF, NLMF, and CLAHE to meet the challenges arising from data deficiency and complexity.
- We analyze the performance of the utilized preprocessing enhancement scheme to demonstrate its role in enhancing the discrimination capability of the proposed model.

The rest of this paper is organized as follows: Section (2) illustrates the phases of the proposed method. Section (3) highlights the experimental results. Section (4) discusses these results. The conclusion of this study is presented in the last section.

## PROPOSED METHOD

In this section, we briefly describe the scenario of the methodology used to achieve the purpose of this study. The proposed method is depicted in Figure 2, which generally consists of two phases: (a) image pre-processing, to overcome the existing drawbacks mentioned in the previous section; (b) training and testing dedicated to image classification.

**Figure 2.**
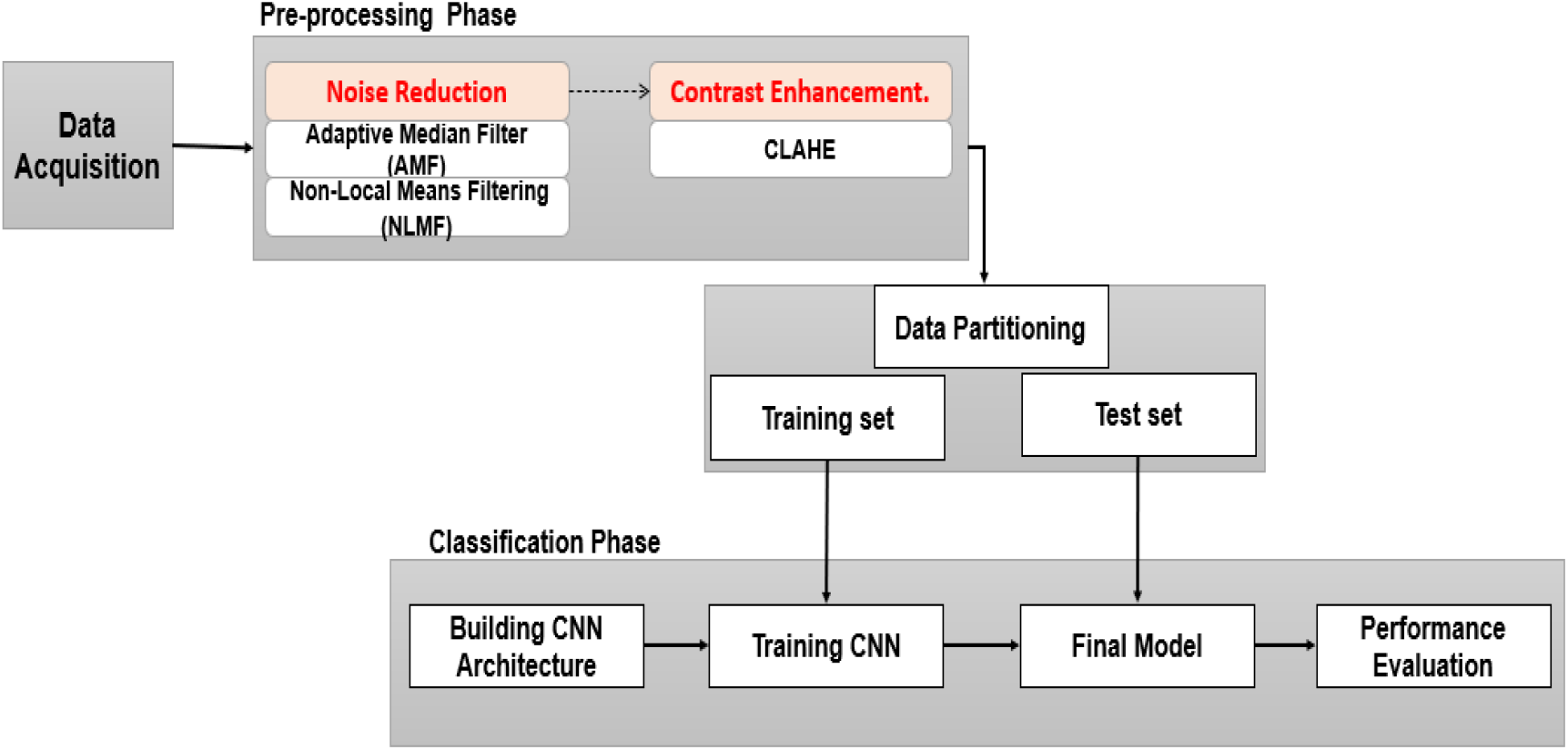
Study Framework.

### Data Acquisition

In this paper, we used the COVIDx dataset used in (Wang et al., 2020) to train and evaluate the proposed model. In brief, COVIDx dataset is an open-source dataset that can be downloaded from https://github.com/lindawangg/COVID-Net/blob/master/docs/COVIDx.md. The instructions given by COVID-Net (Wang et al., 2020) were followed to set up the new dataset. Since the number of X-ray images available for positive Covid-19 cases is very small, more Covid-19 X-ray images from https://github.com/ml-workgroup/covid-19-image-repository as well as https://github.com/armiro/COVID-CXNet/tree/master/chest_xray_images/covid19 were also downloaded to overcome this limitation. Duplicated images were omitted from the new dataset to ensure the proposed model in training is more accurate. So, the actual number of images in the Covid-19 class became 2128 instead of 1770 in COVIDX (updated on 28 January 2021). We used the same test set that was used for evaluation in (Wang et al., 2020), making a slight change by increasing the number of Covid-19 images to 100 images instead of 92. Table 1 summarizes the number of images in each class and the total number of images used for training and testing.

**Table 1.** The number of images for each class.

| Data | Covid19 | Normal | Pneumonia | Total |
| --- | --- | --- | --- | --- |
| train | 2028 | 7181 | 4981 | 14100 |
| test | 100 | 885 | 594 | 1579 |

### Data Pre-processing Method

In this study, we attempt to provide an algorithm that would increase the image quality by using a hybrid technique consisting of noise reduction and contrast enhancement. Specifically, two efficient filters are used for noise reduction while CLAHE is used for contrast enhancement. The first filter is the Adaptive Median Filter (AMF) that removes impulse noise (Ning et al., 2009; Khare and Chugh, 2014). This filter is followed by the Non-Local Means Filtering (NLMF) algorithm that calculates similarity based on patches instead of pixels. Given a discrete noisy image *u* = *u*(*i*) for a pixel *I*, the estimated value of *NL*[*u*](*i*) is measured as the weighted average of all the pixels, i.e.:

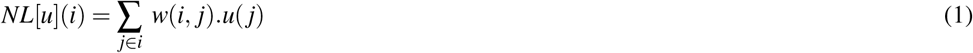

where the weights family *w*(*i, j*) *j* depends on the similarity between the pixels *i* and *j*.

The similarity between the two pixels *i* and *j* is defined by the similarity of the intensity of gray-level vectors *u*(*N*_*i*_) and *u*(*N*_*j*_), where *N*_*l*_ signifies a square neighborhood of fixed size and centered at a pixel *L*. The similarity is measured as a function to minimize the weighted Euclidean distance, 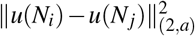 where *a >* 0 is the Gaussian kernel standard deviation. The pixels with a similar gray-level neighborhood to *u*(*N*_*i*_) have larger weights in the average. These weights are defined as,

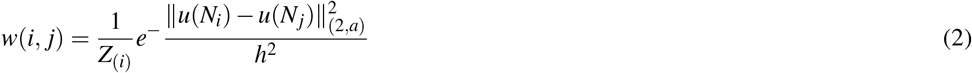

where *Z*_(*i*)_ is the normalizing constant, and the settings *h* works as a filtering degree.

Next, CLAHE is applied to the denoised images to achieve an acceptable visualization and to compensate for the effect of the filtration that may contribute some blurring to the images (Huang et al., 2016; Senthilkumar and Senthilmurugan, 2014).

### Classification Neural Network Model

We used a deep neural network structure called a MobileNet neural network (Howard et al., 2017). Before we pass the input to the neural network, we resized all images to 224 × 224 × 3. These were used as input to the network. Figure 3 depicts the model diagram of the network. The first layer is a Depthwise Conv2D layer of size 3 × 3 × 3 with stride 1, followed by a Conv2D layer with 64 kernels size of 1 × 1 × 3 and stride 1. After that, there is another Depthwise Conv2D layer size of 3 × 3 × 64 with stride 2. Then comes a convolutional layer with 128 kernels and the size of 1 × 1 × 64 with stride 1 followed by a Depthwise separable convolution layer of size 3 × 3 × 128 with stride 1. Then there is another convolution layer with 128 kernels size of 1 × 1 × 128 with stride 1 followed by a Depthwise separable convolution layer of size 3 × 3 × 128 with stride 2. After that, another convolutional layer with 256 kernels and the size of 1 × 1 × 128 with stride 1 is followed by a Depthwise Separable Convolution layer of size 3 × 3 × 256 with stride 2. Then another convolutional layer has 256 kernels with the size of 1 × 1 × 256 with stride 1, followed by Depthwise Separable Convolution layers of sizes of 3 × 3 × 256 with stride 2. Then comes a convolutional layer with 512 kernels with the size of 1 × 1 × 256 with stride 1 followed by another Depthwise separable convolution layers of sizes of 3 × 3 × 512 with stride 1. This is followed by five blocks of layers, each block consisting of convolutional layers with 512 kernels and the size of 1 × 1 × 512 with stride 1 followed by Depthwise separable convolution layers of sizes of 3 × 3 × 512 with stride 1. After that comes another convolutional layer with 1024 kernels of size 1 × 1 × 512, then Depthwise separable convolution layers of sizes of 3 × 3 × 1024 with stride 1. Then again, another convolutional layer with 1024 kernels with the size of 1 × 1 × 1024 with stride 1. Then a dropout layer with the rate of 0.001 is added. The dropout layer’s output goes to two fully connected layers that generate the output of size 128. One fully connected layer is used to predict the mean *µ*, and the other is used to predict the standard deviation *σ* of a Gaussian distribution. The mean *µ* and standard deviation *σ* of a Gaussian distribution are used to calculate the KL loss function. The output of the fully connected layer used to predict the mean *µ* goes to the last layer, a fully connected layer containing the SoftMax activation function that can be used as a classifier, as defined in equation 3, where *v* indicates the output vector, *o* indicates the Objective vector, and *p j* indicates the input to the neuron *j*.

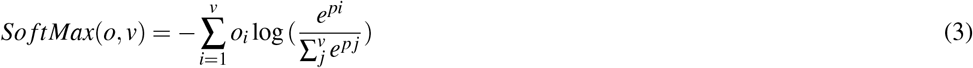

**Figure 3.**
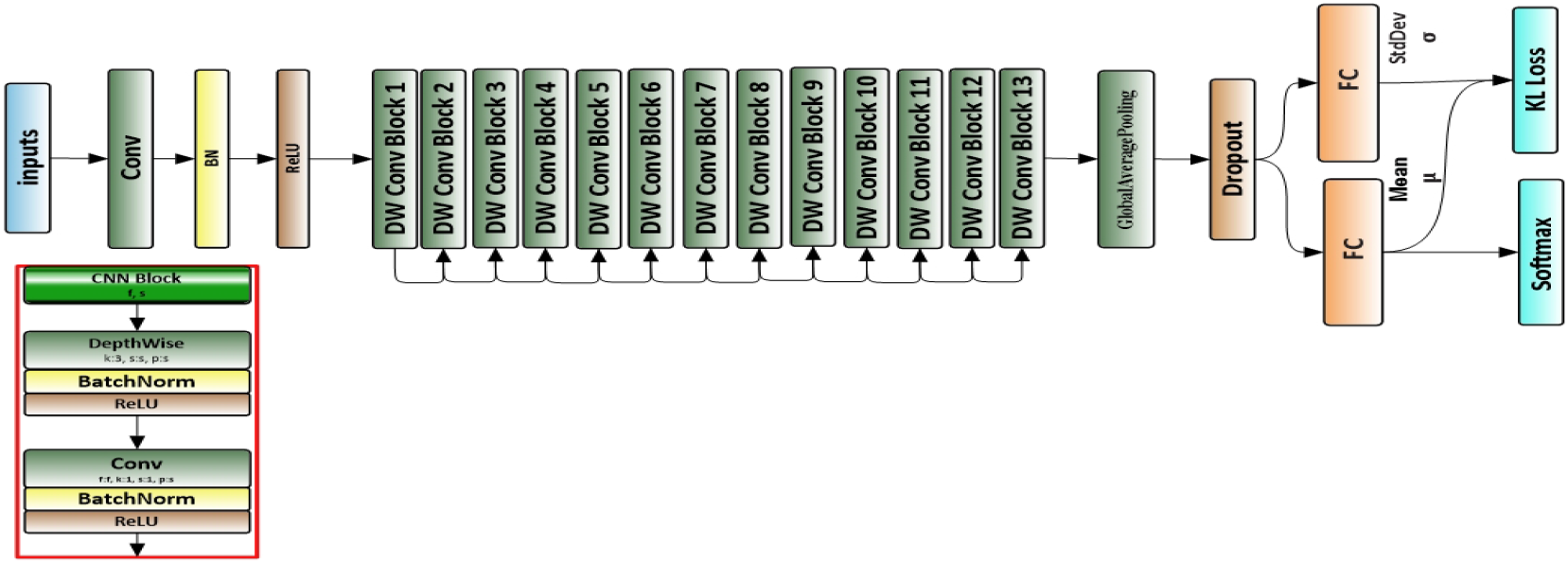
The architecture of the proposed neural network.

The KL divergence between the *µ, σ* distribution and the prior are considered as a regularization which helps to overcome the over-fitting problem defined in equation 4.

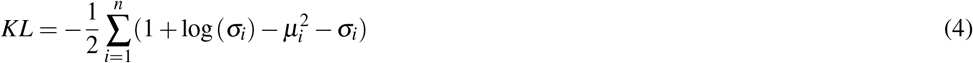

where *n* is the output vector of the average pooling layer with the size of 1024, *µ* is the mean that has been predicted from one fully connected layer, and *σ* is the standard deviation of a Gaussian distribution predicted from one fully connected layer in the network.

As soon as the data is pre-processed, the network is trained with a SoftMax classifier for 200 epochs using Adam optimizer (Kingma and Ba, 2014) on a GPU. The dataset used for training is divided: 70% as a training set and 30% as a validation set. The total number of parameters is 3,488,426, where the number of trainable parameters is 3,466,660, and the non-trainable parameters are 21,766.

### Experiments

All CXRs were resized to the same dimension of 224 × 224 in .jpg format. In the first phase, the AMF window size was taken to be 5 × 5 for effective filtering. The resultant image was subjected to the NLMF technique. The performance of NLMF depends on 7 × 7 of the search window, 5 × 5 of the similarity window, and the degree of filtering *h* = 1. Furthermore, we increased contrast using CLAHE with the bin of 256 and block-size of 128 in slope 3 to get the enhanced images. The proposed model (KL-MOB) was trained using the Python programming language. All experiments were conducted with a Tesla K80 GPU graphics card on Google Collaboratory with Windows 10 operating system. The original and enhanced images were used separately to train the KL-MOB, using Adam optimizer, with the initial learning rate set to 0.00001, on 200 epochs. We passed the images to KL-MOB as the input to predict the CXR image, whether Covid1-9, normal, or pneumonia. Because many functions are not built-in functions in deep learning libraries, such as the relu6 activation function with a max value of 6, we needed to build an interface for the evaluation process that contains all layers in the network as in a training network but which is not used for training. Rather, it is just used to pass on the input image to produce the output result.

### Performance Evaluation

#### Pre-processing Performance Evaluation

The performance of the proposed preprocessing technique was quantified by using various evaluation metrics such as Mean Average Error (MAE) and Peak Signal to Noise Ratio (PSNR). These metrics are desirable since they are fast to quantify.

Definition: *x*(*i, j*) denotes the samples of the original image, *y*(*i, j*) denotes the samples of the output image.*M* and *N* are the number of pixels in row and column directions, respectively. *MAE* is calculated as in equation 5, where a large value means that the images are of poor quality.

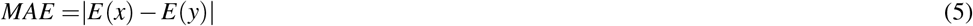

The limited value *PSNR* implies that the images are of low quality. PSNR is described in terms of Mean Square Error *MSE* as follows:

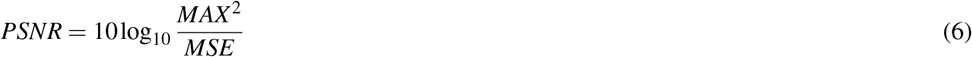

where *MAX* ^2^ is the maximum possible pixel intensity value 255 when the pixel is represented by 8 bits.

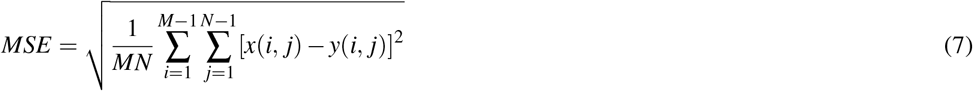

#### Neural Network Performance Evaluation

The test set described in the previous section was used to evaluate KL-MOB. The classification outcome has four cases: True Positive (TP), False Positive (FP), True Negative (TN), and False Negative (FN). The metrics used to measure the performance are Accuracy (ACC), Sensitivity (TPR), Specificity (SPC), and Precision (PPV), defined as follows:

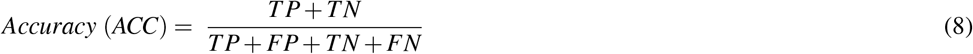

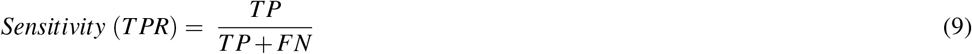

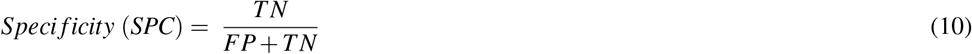

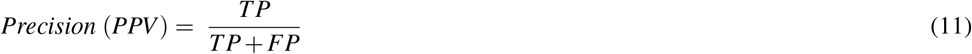

The graph plotted between True Positive Rate (TPR) and False Positive Rate (FPR) is the receiver operating characteristic (ROC) curve. FPR is calculated as follows:

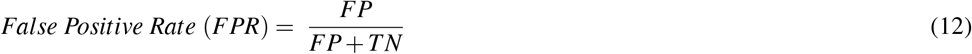

## RESULTS

In the experiments, noise reduction and contrast enhancement performance were evaluated independently since they are two separate issues. The average value was computed for all images in each class. The evaluation results are shown in Tables 2, and 3 for noise reduction and image enhancement, respectively. Figure 4 shows noise reduction techniques that were applied to the original image and the hybrid method used in this work. Though the denoising filters could present smoothing and blurring to the resulting images, this can be enhanced by improving the images’ edges and by highlighting the high-frequency components to remove the residual noise. Figure 5 displays the original images and their enhanced versions

**Table 2.** The average values of PSNR (dB) and MAE for the different noise reduction methods.

| Method | Covid19 |  | Normal |  | Pneumonia |  |
| --- | --- | --- | --- | --- | --- | --- |
|  | PSNR | MAE | PSNR | MAE | PSNR | MAE |
| AMF | 21.91 | 14.46 | 21.19 | 17.88 | 20.43 | 19.47 |
| NLMF | 20.47 | 19.19 | 20.41 | 19.41 | 20.40 | 19.40 |
| our method | 22.04 | 14.38 | 21.21 | 17.59 | 20.45 | 19.32 |

**Table 3.** The average values of PSNR (dB) and MAE for the different contrast enhancement methods.

| Method | Covid19 |  | Normal |  | Pneumonia |  |
| --- | --- | --- | --- | --- | --- | --- |
|  | PSNR | MAE | PSNE | MAE | PSNR | MAE |
| CLAHE | 17.83 | 27.35 | 17.12 | 25.98 | 21.91 | 16.20 |
| our method | 19.14 | 23.13 | 17.28 | 25.45 | 22.11 | 16.01 |

**Figure 4.**
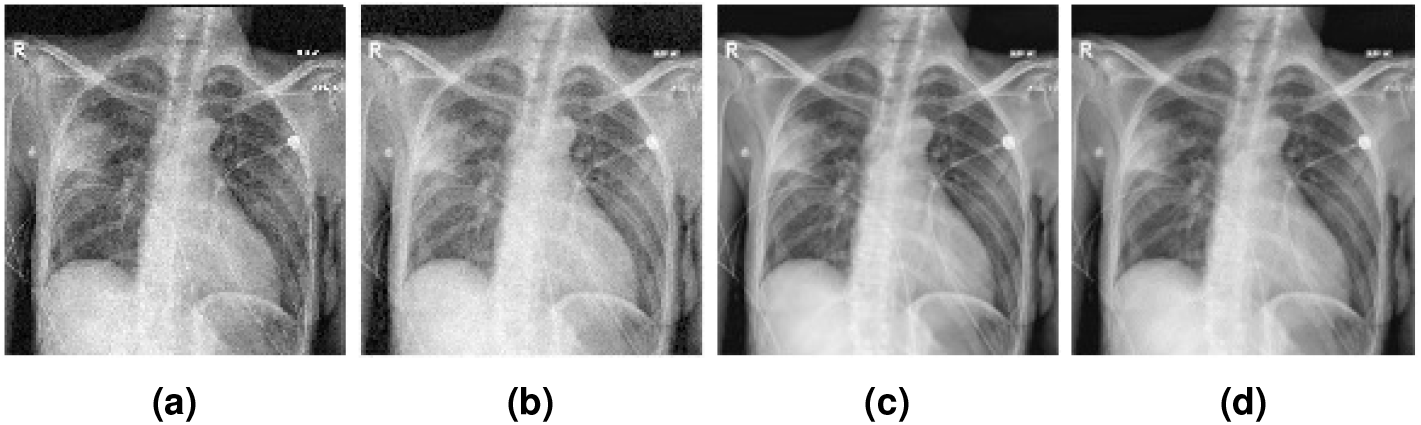
The applied noise reduction techniques on the images: (a) original image, (b) image denoised by AMF, (c) image denoised by NLMF, (d) image denoised by our method.

**Figure 5.**
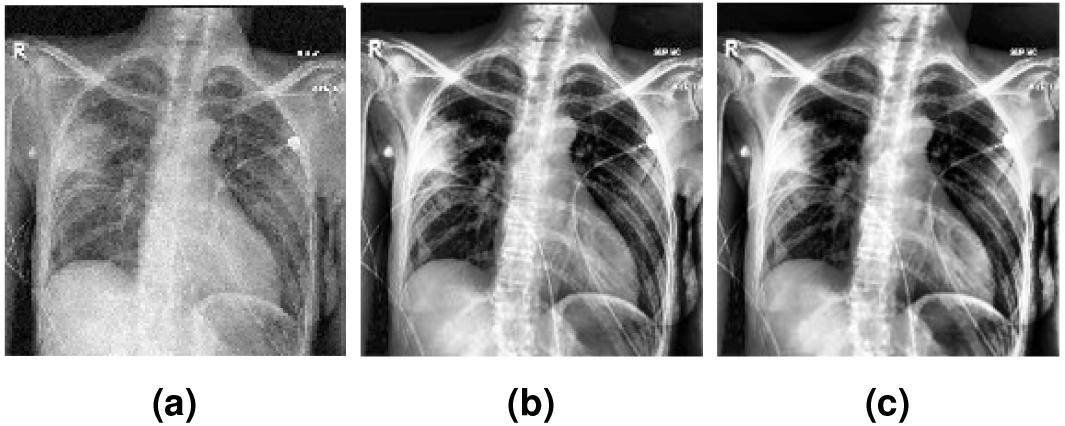
The enhanced image results: (a) original image, (b) image with CLAHE, (c) image enhanced by our method.

In this experiment, the proposed KL-MOB model was trained on original and enhanced images to detect whether they were Covid-19, normal, or pneumonia cases. The evaluation process of the proposed KL-MOB performance was applied to each class of the dataset separately. The comparative performances of KL-MOB for the classification problem on original and enhanced images are shown in Table 4. It is noted that the proposed method has boosted the performance of KL-MOB in Covid-19 detection, as shown in the Figures 6, and 7.

**Table 4.**
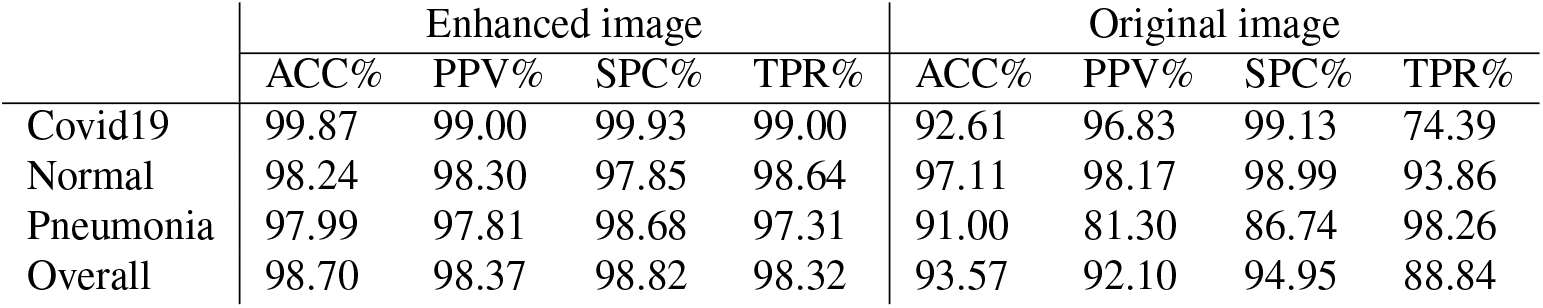
The evaluation metrics of KL-MOB on enhanced and original images.

**Figure 6.**
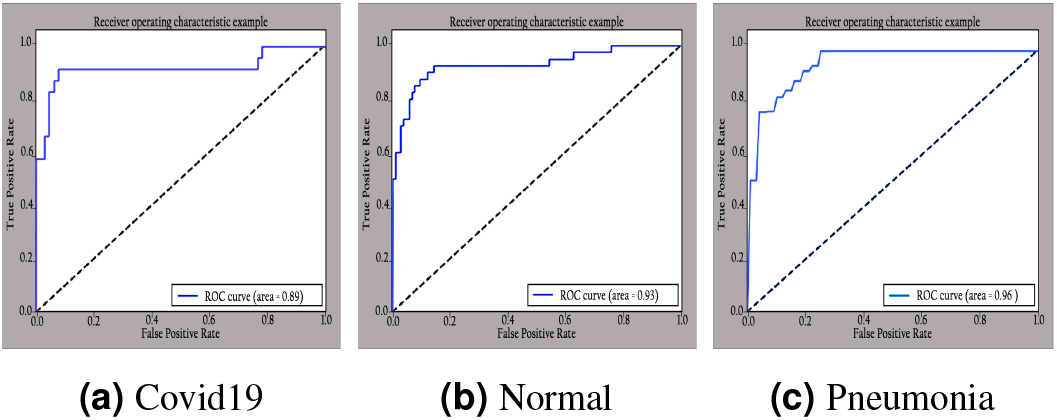
ROC curve of different classes on original images.

**Figure 7.**
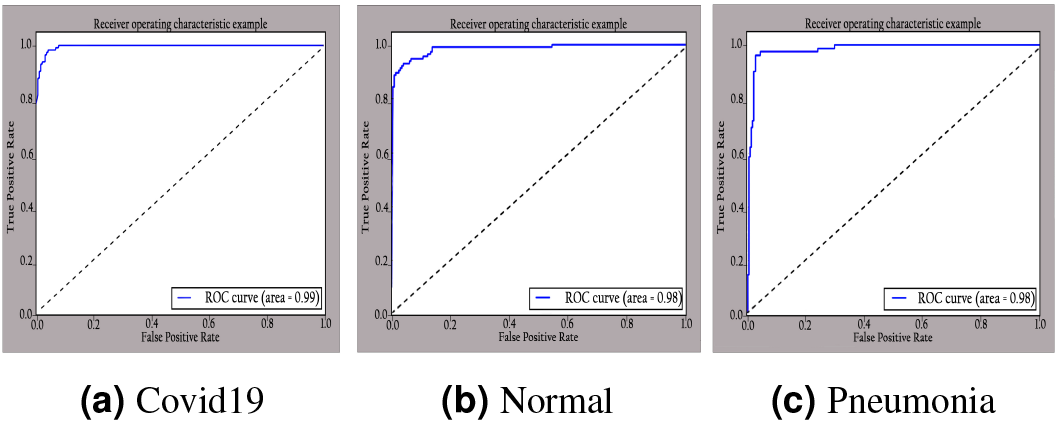
ROC curve of different classes on enhanced images.

## DISCUSSION

In this work, a new approach based on blending noise-eliminate algorithms with contrast enhancement was presented. Adaptation of such an approach introduced a type of hybrid filtering and contrast enhancement for the data set of images used for Covid19 detection. Well-known measurable methods—Peak Signal to Noise Ratio (PSNR) and Mean Average Error (MAE)— were used as Image Quality Measurements (IQM) for assessing and comparing image quality. The results of Table 2 show that using an AMF followed by NLMF was entirely favorable for eliminating noises. Our hybrid algorithm was applied to the entire image instead of parts of the image while preserving important details. Figure 8 illustrates the difference between the original and enhanced CXRs by the method used in this work. Furthermore, we found that the damage of the lung in the enhanced image is more perspicuous than in the original image. In addition, CLAHE with a bin of 256 gave the best PSNR value, as shown in Table 3.

**Figure 8.**
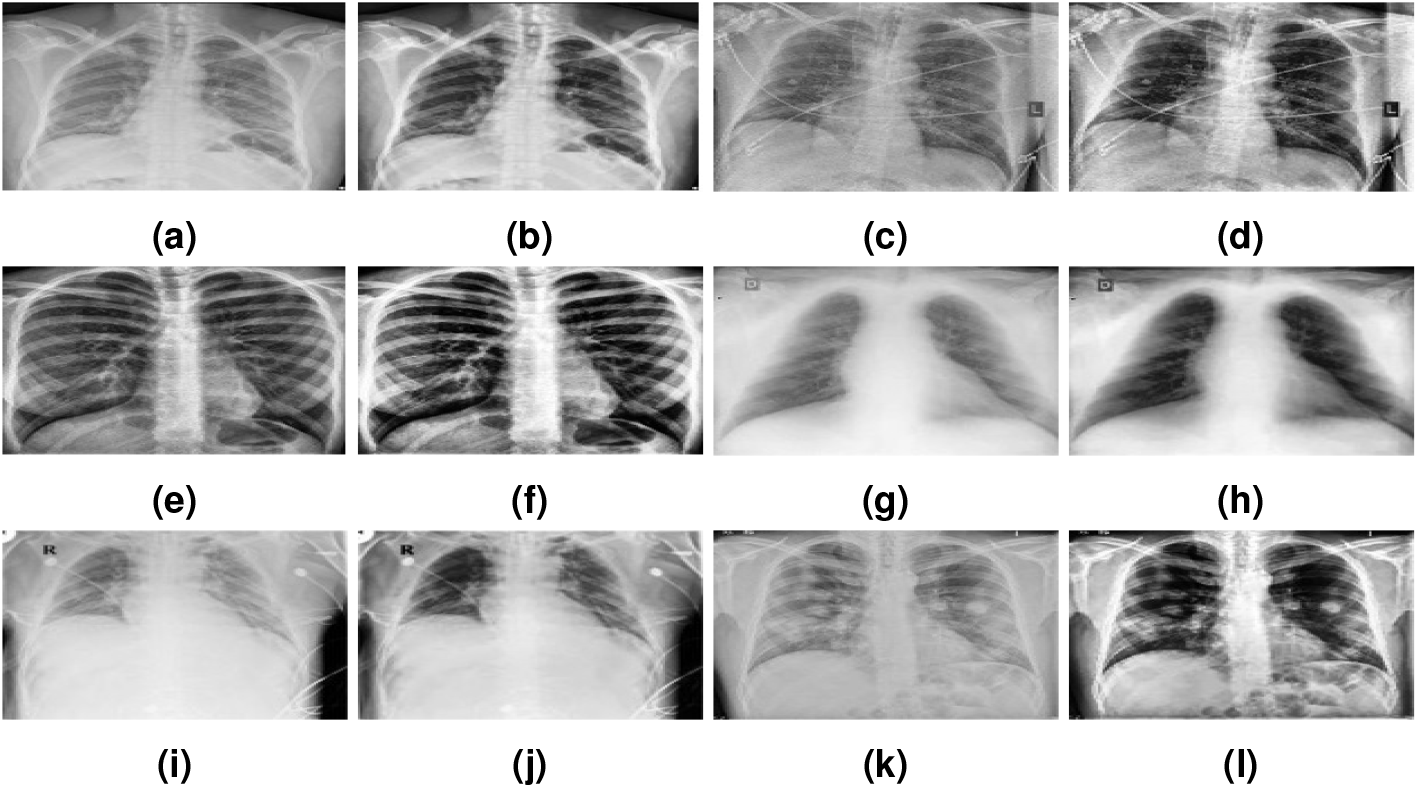
The images in the first and third columns show three cases of original images; the images in the second and fourth columns show three cases of enhanced images.

The results presented in Table 4 show that the proposed network performed well on the test set, which proves that the method used for image pre-processing boosted the performance of KL-MOB. The confusion matrix of our proposed network is depicted in Figure 9. It shows that all classes are identified with high true positives. It is to be noted that the Covid-19 cases are 99% correctly classified by the KL-MOB. There are 1% of Covid-19 cases misclassified as pneumonia (non-Covid-19), and 1.4% of the normal cases are misclassified as pneumonia. Only 0.2% of pneumonia (non-Covid-19) cases are wrongly classified as Covid-19. These results demonstrate that our proposed KL-MOB has good potential in detecting Covid-19; in particular, with limited Covid-19 cases, we show that there is no confusion between the normal and Covid-19 patient groups.

**Figure 9.**
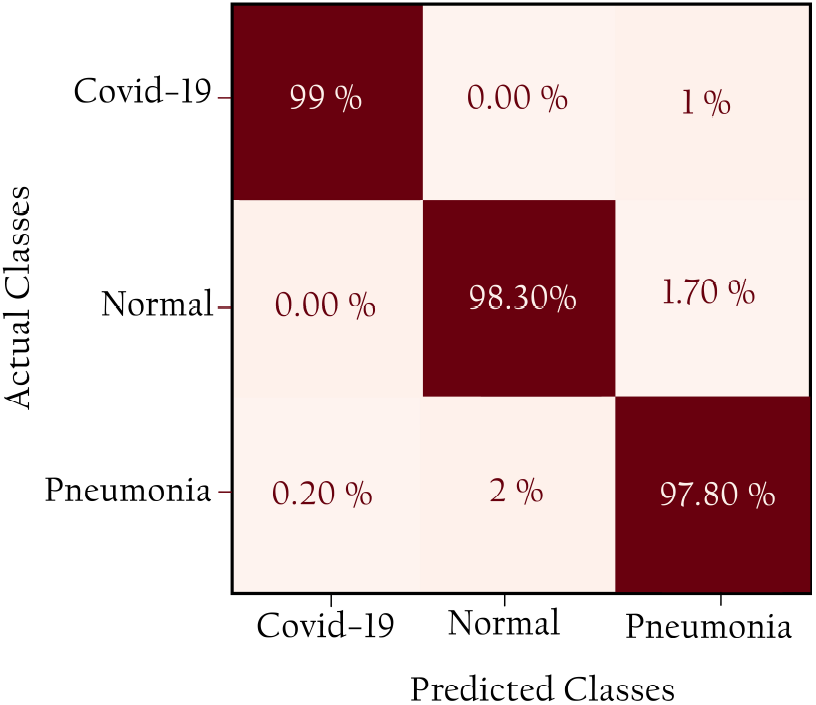
Confusion matrix for KL-MOB on the COVIDx test dataset.

In our experiment range of 100 patient samples of Covid-19, only one was misclassified with a 99.0% PPV for Covid-19, which is an appropriate value compared with 98.9%, and 96.12% for (Wang et al., 2020; Rezaul Karim et al., 2020), respectively. In addition, the results obtained from KL-MOB have been compared with previous studies that used the same or similar datasets for evaluation, as outlined in Table 5. The other studies (Farooq and Hafeez, 2020; Afshar et al., 2020; Hirano et al., 2020; Ucar and Korkmaz, 2020), not included in Table 5 for performance comparisons, utilized smaller datasets. The results showed that KL-MOB is superior to (Wang et al., 2020; Ahmed et al., 2020) across all performance metrics of accuracy, sensitivity (TPR), specificity, and PPV for overall detection.

**Table 5.** Comparative performance of different models.

| Study | Classifier | ACC% | SPC% | TPR% | PPV% |
| --- | --- | --- | --- | --- | --- |
| Wang et al. (2020) | COVID-Net (large) | 95.56 | 96.67 | 93.33 | 93.55 |
| Ahmed et al. (2020) | ReCoNet | 97.48 | 97.39 | 97.53 | 96.27 |
| Rezaul Karim et al. (2020) | DeepCOVIDExplainer | 98.11 | 98.19 | 95.06 | 96.84 |
| proposed method | KL-MOB | 98.7 | 98.82 | 98.32 | 98.37 |

The promising deep learning models used for the detection of Covid from radiography images indicate that deep learning likely still has untapped potential and can possibly play a more significant role in fighting this pandemic. There is definitely still room for improvement, through other processes such as increasing the number of images, implementing another pre-processing technique i.e., data augmentation, utilizing different noise filters, and enhancement techniques.

## CONCLUSION

In this work, we proposed a novel CNN-based MobileNet structure neural network for Covid19 detection using COVIDx, the most widely used public dataset of CXR images to date. As well, the evaluation results show that our approach outperforming a recent approach with accuracy, specificity, sensitivity, and precision of 98.7%, 98.82%, 98.32%, and 98.37%, respectively. The proposed method relied on image manipulation by applying a hybrid technique to enhance the visibility of CXR images. This advanced pre-processing technique made the task of KL-MOB easier and better able to extract features, as it helped to recognize complex patterns from medical images at a level comparable to that of experienced radiologists. The KL loss function was used to boost the performance of KL-MOB which outperformed recent approaches as shown in the obtained results. Considering several essential factors such as Covid-19 infection spreading patterns, image acquisition time, scanner availability, and costs, we hope our findings will be a useful contribution to the fight against Covid-19 and towards an increasing acceptance and adoption of AI-assisted applications in clinical practice. As future work, we will further enhance our method’s performance by including the lateral view of CXR images in our training data, as in some of the cases, frontal view of CXR images does not give a clear idea in diagnosing pneumonia cases. Further, since only a limited amount of CXR images for Covid-19 infection cases, the potential for issues to arise is out-of-distribution is possible, therefore, more unseen data from related distributions is needed for further evaluation. Finally, the enhancement of the images must be verified with a radiologist, which we have not yet been able to do due to the emerging conditions.

## Data Availability

we used the COVIDx dataset used in (Wang et al., 2020) to train and evaluate the proposed model. In brief, the COVIDx dataset is an open-source dataset that can be downloaded from Github.

https://github.com/ml-workgroup/covid-19-image-repository

https://github.com/lindawangg/COVID-Net/blob/master/docs/COVIDx.md

https://github.com/armiro/COVID-CXNet/tree/master/chest_xray_images/covid19

## ACKNOWLEDGMENTS

This work was supported in part by the National Natural Science Foundation [61572177].

